# Estimating the Efficacy of Quarantine and Traffic Blockage for the Epidemic Caused by 2019-nCoV (COVID-19):A Simulation Analysis

**DOI:** 10.1101/2020.02.14.20022913

**Authors:** Deqiang Li, Zhicheng Liu, Qinghe Liu, Zefei Gao, Junkai Zhu, Junyan Yang, Qiao Wang

## Abstract

**Background:** Since the 2019-nCoV (COVID-19) outbreaks in Wuhan, China, the cumulative number of confirmed cases is increasing every day, and a large number of populations all over the world are at risk. The quarantine and traffic blockage can alleviate the risk of the epidemic and the infections, henceforth evaluating the efficacy of such actions is essential to inform policy makers and raise the public awareness of the importance of self-isolation and quarantine.

**Method:** We collected confirmed case data and the migration data, and introduced the quarantine factor and traffic blockage factor to the Flow-SEIR model. By varying the quarantine factor and traffic blockage factor, we simulated the change of the peak number and arrival time of infections, then the efficacy of these two intervation measures can be analyzed in our simulation. In our study, the self-protection at home is also included in quarantine.

**Results:** In the simulated results, the quarantine and traffic blockage are effective for epidemic control. For Hubei province, the current quarantine factor is estimaed to be 0.405, which means around 40.5% of suceptibles who are close contacting with are in quarantine, and the current traffic blockage factor is estimaed to be 0.66, which indicates around 34% of suceptibles who had flowed out from Hubei. For the other provinces outside Hubei, the current quarantine factor is estimated to be 0.285, and the current traffic blockage factor is estimated to be 0.26. With the quarantine and traffic blockage factor increasing, the number of infections decrease dramatically. We also simulated the start dates of quarantine and traffic blockage at four time points, the simulated results show that the early of warning is also effective for epidemic containing. However, provincial level traffic blockage can only alleviate 21.06% - 22.38% of the peak number of infections. In general, the quarantine is much more effective than the traffic blockage control.

**Conclusion:** Both of quarantine and traffic blockage are effective ways to control the spread of COVID-19. However, the eff icacy of quarantine is found to be much stronger than that of traffic blockage. Considering traffic blockage may also cause huge losses of economy, we propose to gradually deregulate the traffic blockage, and improve quarantine instead. Also, there might be a large number of asymptomatic carriers of COVID-19, the quarantine should be continued for a long time until the epidemic is totally under control.

## Background

Since the 2019-nCoV (COVID-19) outbreaks in Wuhan, China, the cumulative number of confirmed cases is increasing every day, and a large number of populations all over the world are at risk. The recent study [1-4] on the efficacy of traffic blockage for the COVID-19 indicated that the the population flows will certainly increase the cumulative number of cases and the quarantine and traffic blockage will lower this risk.

On January 24, 2020, J. M. Read *et al* [5] applied the transmission model and found that the reproduction number of infection is 3.8, and 72% - 75% of the transmission needed to be restricted in order to inhibit the growing infection rate; In addition, they found that traffic blockage only slowed the spread of the epidemic by 24.9%.

On January 27, 2020, B. Tang *et al* [6] proposed a deterministic compartmental model based on clinical progression of the epidemic. They showed that the reproduction number of infection is 6.47 and the intervention measures can reduce the reproductive number of infection and the risk of transmission. The sensitivity results show that the number of infections in Beijing will decrease by 91.14% in 7 days with the intervation of travel.

On Feb.4, 2020, S. Ai *et al* [7] investigated both the effectiveness of closure of Wuhan and that of restricting the flow of people in and out the provinces (cities) in China.

On Feb.9, 2020, X. Li *et al* [8] simulated the dynamics of the 2019-nCOV (COVID-19), on account of the strict control measures in Wuhan and Beijing. They showed that after the the traffic blockage on Jan.23,2020, the reproductive number in Wuhan will be 2.5, significantly lower than before.

Unlike our previous work [9] on H7N9 avian influenza which was a retrospective analysis, we are now doing predictive analysis of COVID-19 in an emergency. Beside our Flow-SEIHR model based predition for the trends of COVID-19 [10], this paper will focus on quantifying the quarantine factor and the traffic blockage factor such that we may numerically evaluate the effects of various policies. Specifically, we apply a Flow-SEIR model, a simplified version of Flow-SEIHR model proposed in [10], which may enable us to perform a wide range of assessments nationwide.

## Methods

The outbreaks of COVID-19 occured around the Chinese Spring Festival, when a huge number of populatin flows in and out the origin outbreak area, Wuhan, which resulted in that the epidemic was severe in mainland of China and worldwide. The policy of traffic blockage was executed since Janurary 23^rd^, 2020, after then the chance of cross-infection decreased sharply. Here we make the following assumptions regrading to the status quo of the epidemic and the SARS empirical parameters.

A1: The outbreak of COVID-19 started on the beginning of Dec., 2019.

A2: The flows in domestic provinces are concluded, the abroad flow-in is not considered.

A3: Only the susceptible with no clinical symptoms can flow in and out, the exposed and infections will be quarantined.

A4: There is no super spreaders, in other words, the chance of infected was equal to those who are contact with the infections.

A5: The COVID-19 has weak seasonality in its transmission, unlike influenza.

A6: The recovery rate is time-dependent and will linearly increase.

A7: The empirical parameters on SARS will also work on COVID-19.

A8: Most people will follow the official instructions and stay at home after the traffic blockage.

We denote S, E, I, R,F in the flow-SEIR model [10] and the parameters as shown in Table 1. In addition, we introduce the traffic blockage factor τ, and the quarantine factor ε, which stands for the quantifications of traffic blockage and quarantine, respectively. Since the people was urged to stay at home, this self-protection is also regarded as quarantine.

**Table 1.** Description of the variables of the model and control factors

| Variable | Description |
| --- | --- |
| $S$ | Number of those susceptible |
| $E$ | Number of those exposed |
| $I$ | Number of those infective |
| $R$ | Number of those recovered |
| $F$ | Population flows in and out |
| $\beta$ | Rate of those who are susceptible and being infected |
| $\alpha$ | Rate of those who are exposed and being infectious. |
| $\gamma_1$ | Those who are infectious and being recovered. |
| $\gamma_2$ | Those who are exposed and being recovered. |
| $\varepsilon$ | Rate of susceptibles who are in quarantine |
| $\tau$ | Rate of susceptibles who flowed out |

### Flow-SEIR model

Basically, we follow the standard SEIR [11-13] model to evaluate the state of infections. However, since the epidemic of COVID-19 appeared around the Chinese Spring Festival, the nationwide traffic should be modeled by the traffic network flow, in order to characterize the huge number of population flows in and out all provinces (cities), which formed the flow-SEHIR model in [10]. Here we simplify it into a Flow-SEIR model, and then estimate the influence of the migration and the efficacy of traffic blockage. The Flow-SEIR model is:

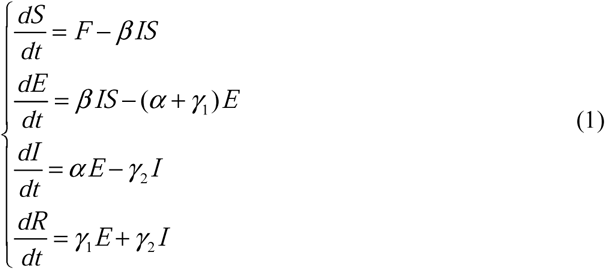

where all the variables are presented in Table 1. The migration data can represent the quantification of the susceptibles flow function *F* in the first equation of (1). In practice, we collect the migration data from the Baidu Migration API, which is available online. More precisely, the susceptibles flow to the *i*th province from other areas can be expressed as follows:

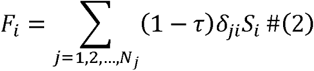

where *δ*_*ji*_ is the ratio of migration population to the whole population independent of the traffic blockage. In equation (2), the traffic blockage factor τis introduced, which ranges from 0 to 1. Here τ tends to 0 implies free, and 1 blockage.

According to our assumptions, the transmission with quarantine can be rewritten as:

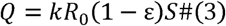

where Q is the number of susceptibles who are independent of quarantine, and *k* is a constant which will be estimated, R_0_ is basic reproductive number [12,13]. As the epidemic grows fastly, the public was aware of cross-infection, so the quarantine factor *ε* will increase to 1, thus is in decline gradually.

In appendix 1, all the parameters concerned in equaration (1) can be estimatied by fitting the confirmed data (cf. [16,17]). The practical considerations are also discussed there in appendix 1.

### Data source

All the data we used is available online. The data of confirmed cases is released by the National Health Commission of China and each provinces’ health commissions, and the migration data is from Baidu Migration API. For data source websites, please see Appendix 2

## Results

### The efficacy of quarantine and traffic blockage

Both the quarantine factors and traffic blockage factors are taken between 0 to 1. In Fig 2, the current quarantine factor of Hubei province is estimaed to be 0.405, which means around 40.5% of suceptibles who are close contacting with are in quarantine. If we increase the factor by 0.01, the peak number of infections will decrease by 5.53%. For the other provinces outside Hubei, the current quarantine factor is estimated to be 0.285, if we incease the factor by 0.01, the peak number of infections will decrease by 14.64%. On the other hand, when *ε* is close to 1, all the population is isolated from others, the peak value will decrease by 89.68%. However, when *ε* tends to 0, all the population flows are control free, and the peak value will increase by 20.4%.

**FIGURE 1.**
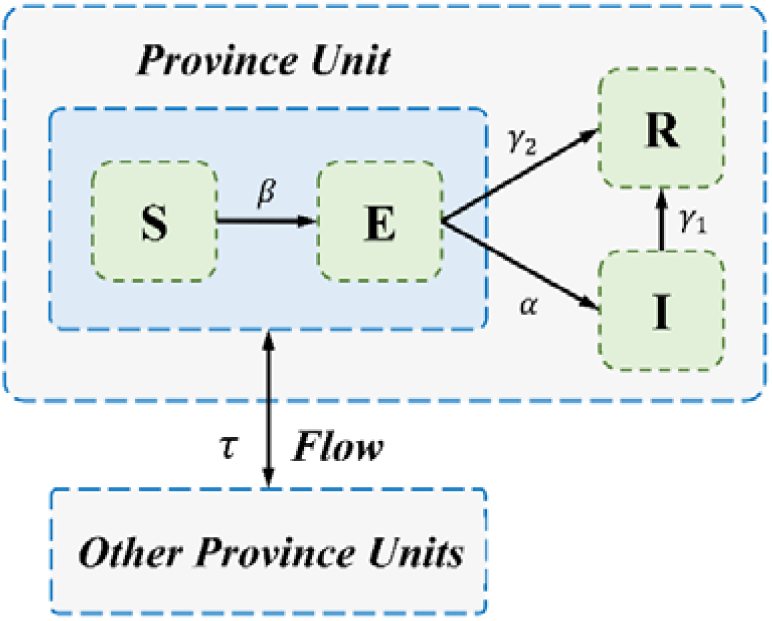
Flow-SEIR model, based on the SEIR model, was proposed to to estimate the epidemic trend with a large number of population flowing in and out all provinces of China during the Chinese Spring Festival. And the efficacy of traffic blockage can be estimated in this model.

**FIGURE 2.**
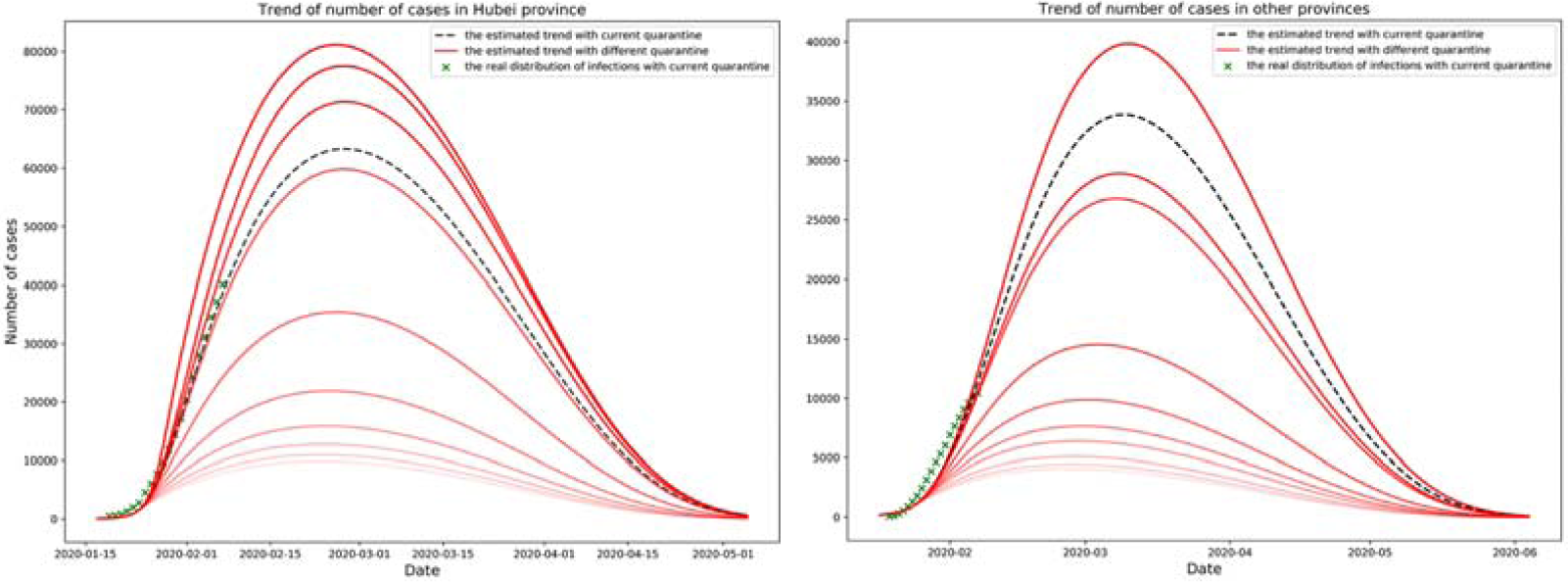
Confirmed cases under different *ε*, the x-axis is date, the y-axis is number of confirmed cases.

As shown in Fig 3, the peak number of cases decrease as the traffic blockage factor increases. For Hubei province, the current traffic blockage factor is estimaed to be 0.66, which indicates around 34% of suceptibles who flowed out from Hubei, if we increase the factor by 0.1, the peak number of infections will decrease by 2.14%. For the provinces except Hubei, the current traffic blockage factor is estimated to be 0.26, if we incease the factor by 0.1, the peak number of infections will decrease by 4.01%. When the traffic blockage factor *τ* is close to 1, the target area is almost completely in blockage, the peak value will decrease by 21.06% - 22.39% to the case that τ tends to 0.

**FIGURE 3.**
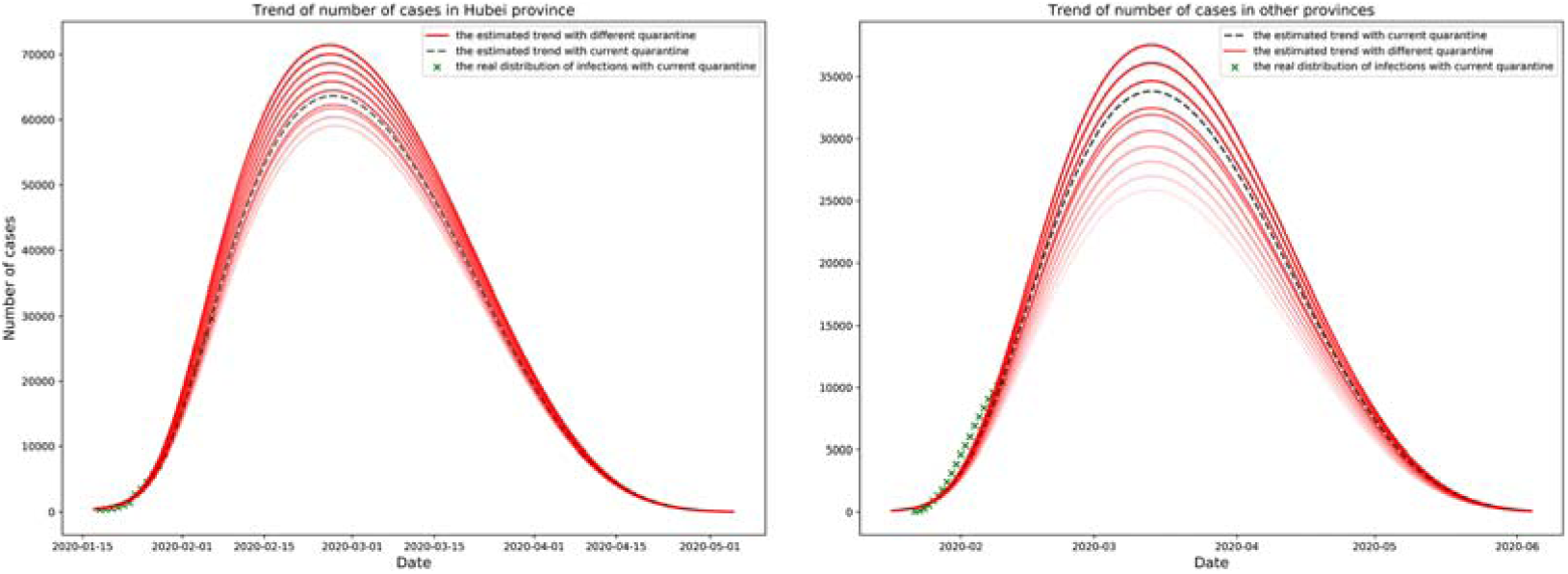
Confirmed cases under different τ, the x-axis is date, the y-axis is the number of confirmed cases.

Clearly, self-isolation and quarantine is more effective than traffic blockage.

### The efficacy of earlier warning of COVID-19

We estimated the trend of confirmed cases with quarantine at different time periods. As shown in Fig 4, the quarantine factor *ε* increase by 0.01, the peak arrival time in Hubei will arrive 0-2 days earlier, and the peak arrival time of other provinces outside Hubei will arrive 1-3 days earlier. For Hubei Province, if the quarantine is 1 week delay, the peak value will increase by 8.50% - 10.95%. Oppsitely, if the quarantine is 1 week earlier, the peak value will decrease by 24.26% - 25.33%, and 2 weeks by 57.10% - 57.46%. For the other provinces, if the quarantine is 1 week of delay, the peak value will increase by 20.92% - 22.77%, but 1 (or 2, resp.) week earlier will decrease by 32.78% - 35.60% (or 63.75% - 67.13%, resp.).

**FIGURE 4.**
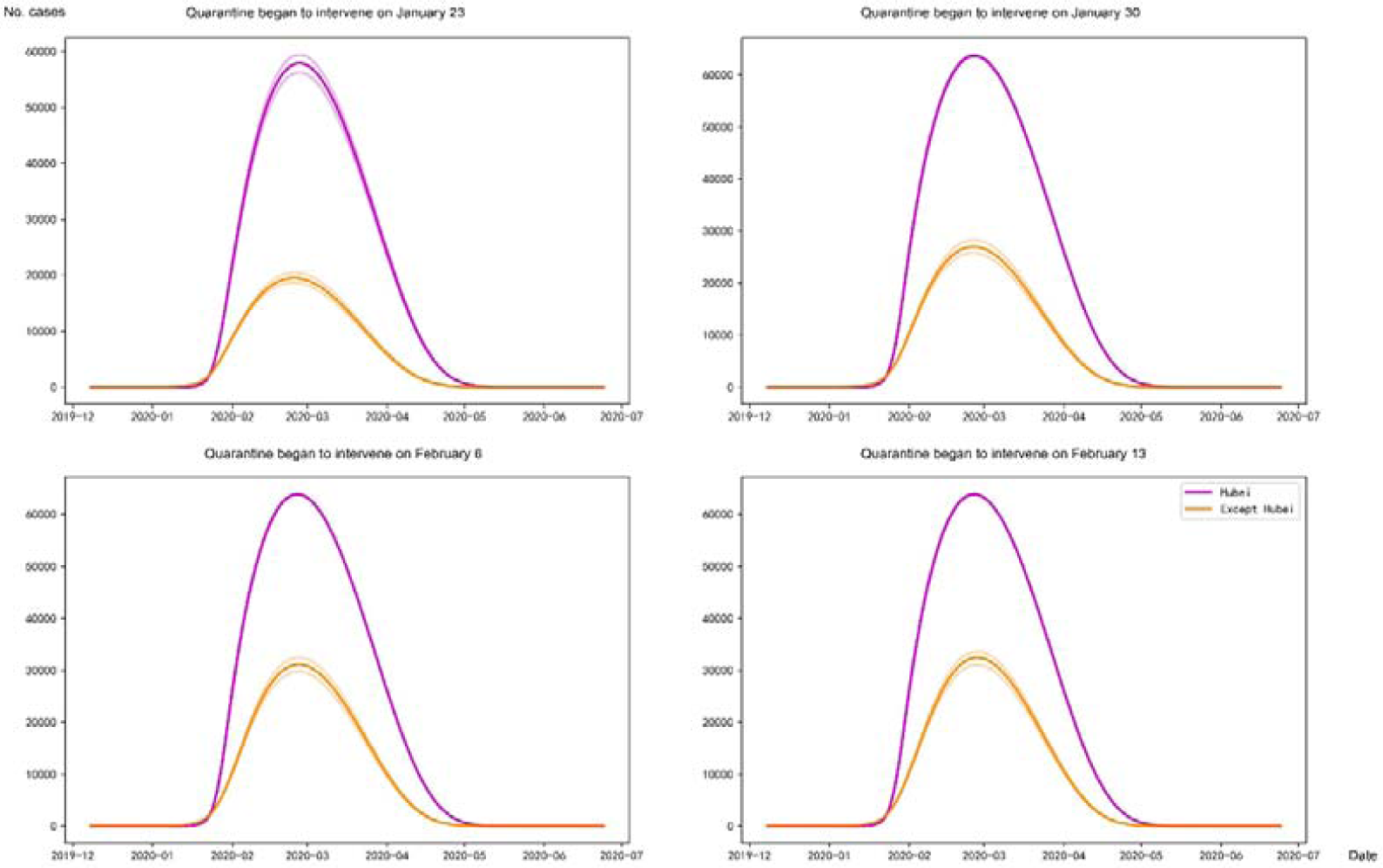
The quarantine factoris *ε* set to take effect on four time: January 23, January 30, Feburary 6, Feburary 13. The x-axis is the date and the y-axis is the number of patients.

Concretely, we estimated the trends of epidemic, assume that the traffic blockage started on Jan. 9, Jan. 16, Jan. 23 and Jan. 30, respectively. In this experiment, we assumed that the traffic flow between provinces would be completely cut off. As shown in Fig 5, when the traffic blockage factor τ increase by 0.1 for Hubei, the peak arrival time will be earlier and the peak number of cases will be lower. If the traffic blockage is one week earlier, the peak number of cases only decreased by 3.10% - 3.79%, and two weeks by 6.64% - 8.08%. This illustrates that in the eve of city closure in Wuhan, the epidemic in Hubei province had been severe, and the transmission rate was quite high. However, in other provinces outside Hubei, if the traffic blockage is one week earlier, the peak number of cases will decrease by 15.30% - 16.70%, and two weeks earlier, it will decrease by 31.74% - 33.09%.

**FIGURE 5.**
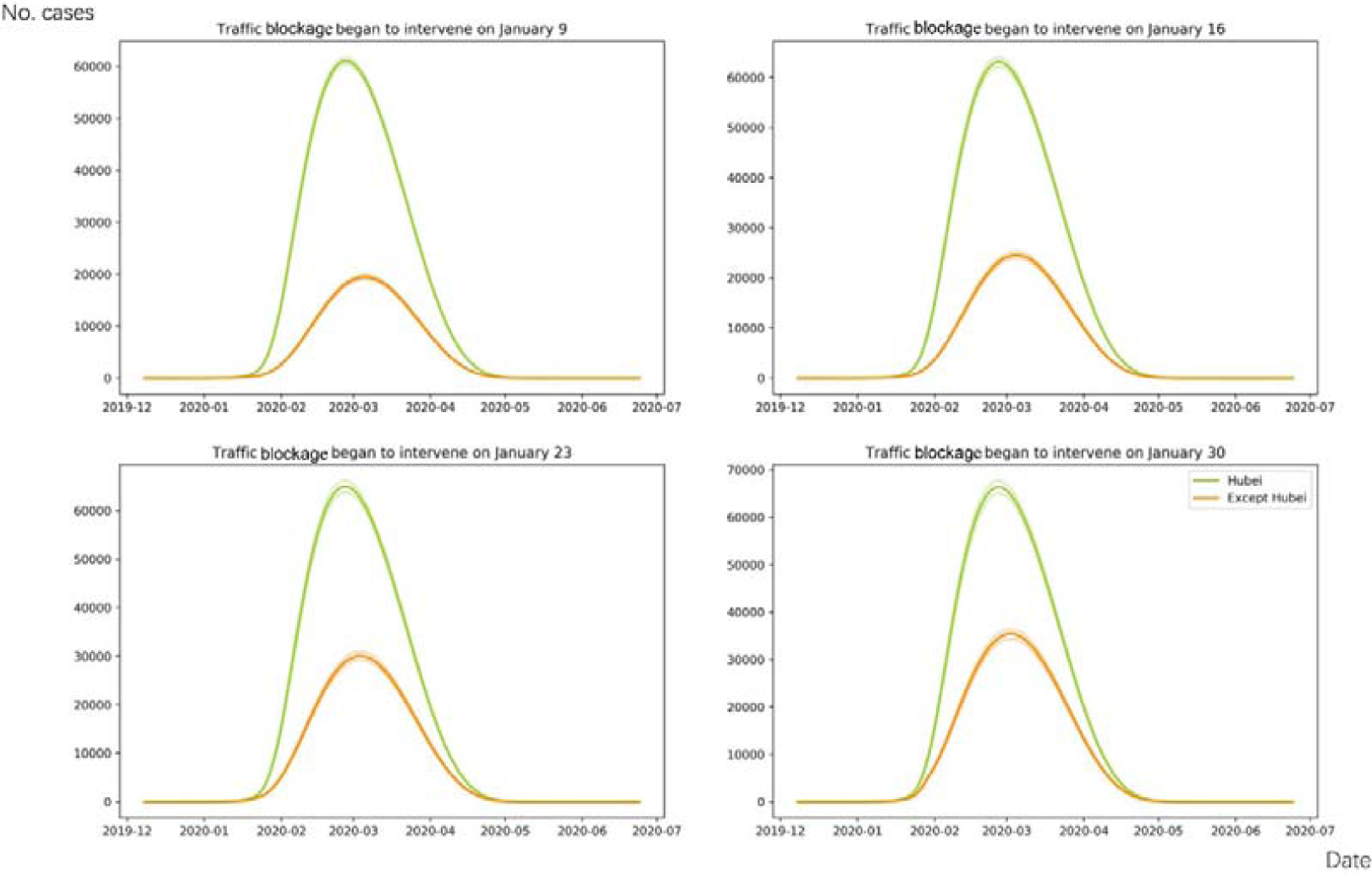
The traffic blockage factor τ is set to take effect on four time: January 9, January 16, January 23, January 30. The x-axis is the date and the y-axis is the number of patients.

Finally, we estimated the daily change of number of cases nationwide. If there is one day earlier of traffic blockage, about 3,600 cases will eventually be reduced in the mainland of China, but one day delay might increase 1,800 cases.

## Conclusions

In this study, we have used a Flow-SEIR model to numerically estimate the efficacy of the quarantine and traffic blockage. We introduced both the the quarantine factor and the traffic blockage factor, then the flow variable and transmission rate in the model will vary as the change of these two intervation factors, then the effectiveness of the traffic blockage and quarantine can be simulated and evaluated.

In the simulated results, we conclude that if the masses take protective measures, the peak number of cases will decrease greatly by 89.68%. However, if the masses completely ignored self-isolation, the peak is predicted to increase by 20.40%. And if the external population flows to the target area is almost completely blocked, the peak number will decrease by 21.06%-22.38%. Earlier warning, timely traffic blockage and quarantine measures are extremely effective, especially for areas with slight epidemic situations. The experimental results also show that if there is one day earlier of traffic blockage, about 3,600 cases will eventually be reduced in the mainland China, and one day delay will result in additional 1,800 cases at risk.

## Discussion

Applying the latest migration population data from Baidu Migration, and varying the traffic blockage factor, we can use the Flow-SEIR model to simulate the number of infections and then intuitively understand the efficacy of the traffic blockage.

However, our study has several major limitations. First, the accuracy of results can be improved provided the source of data is more reliable and sufficient. Since in Hubei the medical resources are overburdened and the methods to diagnose the COVID-9 are not accurate, even the diagnosis changed several times, the underdiagnosis and misdiagnosis are invetiable, then the confirmed cases are underestimated. Secondly, there is heterogenity in transmission, it is not added in our study, and we also did not district the age distribution in our model.

The study has numerically estimated the efficacy of traffic blockage and quarantine. We confirm by simulated quantification evidence that the traffic blockage is effective in controlling the epidemic of COVID-19, and the quarantine is a more effective way to help the public to prevent cross-infection. With the development of the epidemic, we have to admit the fact that there are still a number of population who are exposed to the COVID-19 without clinical symptoms [18-20]. Those population may carry the COVID-19 for several days and might infect others who are closely contacting with, which makes the potentional risk. Thus we encourage the people to be self-protective until the epidemic is totally under control.

## Data Availability

We use the Provincial-Level Migration Data of China from
Baidu migration API, and Official Reported Cases Data. All the data is avaliable from the Internet.

https://www.who.int/health-topics/coronavirus

http://www.nhc.gov.cn/

http://wjw.beijing.gov.cn/

http://qianxi.baidu.com

http://wjw.hubei.gov.cn/

## Abbreviations

2019-nCoV: 2019 Novel Coronavirus
SARS-COV-2: severe acute respiratory syndrome coronavirus 2
Flow-SEIR model: flow-suceptible-exposed-infection-recovery model
COVID-19: corona virus disease 2019.

## APPENDIX 1 Parameters estimation

According to our the assumption A1, firstly we estimated the number of potential suceptibles by the iteration process. We define the loss function as follows:

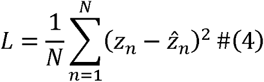

where *N* is the length of the real data, which in practical is the count of days., z_n_ is the *n*th real data value, and *n=1,2,…N*, 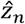 is the *n*th predicted value, based on the model. We fitted the confirmed data from Jan.12 to Feb. 7 to get the optimal number of potential suceptibles by minimizing the loss value. The loss value is small enough to make the estimated results fitting the real data well. The parameters in the Flow-SEIR model can also be estimated in the same way.

For the problem of high death rate in Hubei Province, we reduced the ratio of *γ*_1_ and *γ*_2_ of Hubei’s model according to the real data that *γ*_1*real*_=q_1_*γ*_1,_ *γ*_2real=_ q_2_*γ*_2_ For the situation that the medical resouces are limited in Wuhan, the number of infections will be larger than the released data, we enlarged the ratio of α of Hubei’s model, that α_*real*_=*p*α

And there is data latency problem, because the epidemic has an incubation period and the infections can not be diagnosed timely. For the data latency problem, it can be assumed that the number of confirmed cases published on one day is the number of confirmed cases *n*_*i*_ days ago, and *n*_*i*_ is the number of delayed days in the *i*th province, expressed as

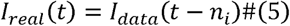

For Hubei, due to the above factors, the latency will be relatively long, which may reach 5-7 days. For other provinces, if patients can get treatment in time, the delay should only include the time waiting for the result of the diagnostic reagents, is about 1-2 day.

## APPENDIX 2 Data Source

### Provincial-Level Migration Data of China

http://qianxi.baidu.com/

### Official Reported Cases Data

https://www.who.int/health-topics/coronavirus

http://www.nhc.gov.cn/

http://wjw.beijing.gov.cn/

http://wjw.fujian.gov.cn/

http://wsjk.tj.gov.cn/

http://wsjkw.hebei.gov.cn/

http://www.xjhfpc.gov.cn/

http://wsjkw.nx.gov.cn/

http://ynswsjkw.yn.gov.cn/wjwWebsite/web/index

http://www.gzhfpc.gov.cn/

http://wsjkw.sc.gov.cn/

http://wsjkw.cq.gov.cn/

http://wst.hainan.gov.cn/swjw/index.html

http://wsjkw.gxzf.gov.cn/

http://wsjkw.gd.gov.cn/

http://wjw.hunan.gov.cn/

http://wjw.hubei.gov.cn/

http://www.hnwsjsw.gov.cn/

http://wsjkw.shandong.gov.cn/

http://hc.jiangxi.gov.cn/

http://wjw.ah.gov.cn/

http://www.zjwjw.gov.cn/

http://wjw.jiangsu.gov.cn/

http://wsjkw.sh.gov.cn/

http://wsjkw.jl.gov.cn/

http://wjw.nmg.gov.cn/

http://wjw.beijing.gov.cn/

http://sxwjw.shaanxi.gov.cn/

http://wjw.shanxi.gov.cn/

http://wsjk.ln.gov.cn/

http://wsjk.gansu.gov.cn/

http://wsjkw.hlj.gov.cn/

https://wsjkw.qinghai.gov.cn/

http://wjw.xizang.gov.cn/

